# Skin Seed Amplification Assay differentiates Multiple System Atrophy from Parkinson’s Disease

**DOI:** 10.64898/2025.12.11.25341627

**Authors:** Anastasia Kuzkina, Diego Rodriguez, Celine Panzer, Alain Ndayisaba, Antonia Kohl, Shreya Rai, Karie Shen, Lisa Harder-Rauschenberger, Olivia Laun, Julia Meehan, Sucika Perumalla, Elena Salabasidou, Kristie Jones, Dalina Ceku, Robert Blum, Susanne Knorr, Camelia M. Monoranu, Claudia Sommer, Jens Volkmann, Barbara K. Changizi, Vikram Khurana, Kathrin Doppler

## Abstract

**Background:** Misdiagnoses of multiple system atrophy (MSA) and Parkinson’s disease (PD) remain common due to overlapping clinical features.

**Objective:** To develop and assess a skin α-synuclein seed amplification assay (αSyn-SAA) for detecting pathological α-synuclein and distinguishing MSA from PD.

**Methods:** In this blinded, cross-sectional study conducted across two laboratories, 308 skin biopsies from 117 participants were analyzed using a standardized αSyn-SAA protocol. The cohort included 42 PD, 31 MSA, 30 controls, and 14 with progressive supranuclear palsy (PSP). Biopsies were obtained from up to four anatomical sites and positivity was defined as ≥75% positive replicates. Kinetic parameters were analyzed to differentiate PD and MSA, and clinical features were correlated with αSyn-SAA positivity. Fibril morphology from amplified skin samples was compared to postmortem brain using proteinase K digestion and electron microscopy.

**Results:** αSyn-SAA detected synucleinopathy in 86% of PD and 77% of MSA patients, with 90% specificity versus controls and 86% versus PSP. ROC analysis showed that maximum fluorescence (Fmax, AUC = 0.93) and lag time (AUC = 0.91) distinguished PD from MSA at the biopsy level. A 50% mean Fmax cutoff across positive samples accurately differentiated synucleinopathies with 90% sensitivity and 85% specificity. αSyn-SAA positivity correlated with RBD in both disorders and with autonomic features in MSA. Amplified fibrils displayed distinct structural and proteolytic profiles between PD and MSA, consistent with brain-derived aggregates.

**Conclusion:** Skin αSyn-SAA detects pathological α-synuclein in living patients with high accuracy and distinguishes MSA from PD based on assay kinetics and fibril strain properties.

## Introduction

Distinguishing Parkinson’s disease (PD) from multiple system atrophy (MSA) remains a major diagnostic challenge. Both are defined by the accumulation of α-synuclein (αSyn) in neurons and glia^1–3^. Overlapping clinical features and the absence of disease-specific biomarkers complicate early diagnosis. Definitive confirmation requires neuropathological identification of characteristic patterns of neurodegeneration and αSyn inclusions^2,3^. Neuroimaging can support the diagnosis of MSA in later stages, but early differentiation from PD remains challenging. Improving early diagnosis is critical to optimize care, enable clinical trial enrollment, and accelerate therapeutic discovery.

Cerebrospinal fluid (CSF) αSyn seed amplification assays (αSyn-SAA) detect misfolded αSyn with high sensitivity and specificity. Several protocols show promise in differentiating PD from MSA^4–6^. However, not all CSF-based assays reliably distinguish, and some fail to detect αSyn seeding activity in MSA altogether^7,8^. Moreover, lumbar punctures are invasive and can be challenging in routine clinical settings. Misfolded αSyn pathology is present in peripheral tissues, including skin^9–14^, saliva, olfactory mucosa, and blood^15^, prompting interest in developing less invasive biomarkers.

Cutaneous αSyn aggregation, typically detected by pS129 immunostaining (p-αSyn), has been demonstrated in PD, MSA, and dementia with Lewy bodies. It is also present in non-motor synucleinopathies such as REM-sleep behavior disorder (RBD)^16–18^ and pure autonomic failure^19^. In MSA, p-αSyn aggregates predominate in somatosensory nerve fibers, whereas in PD, they are primarily localized to cutaneous autonomic nerve fibers^20–22^. While this technique offers anatomical insight, it does not capture strain-specific αSyn conformations.

By contrast, αSyn-SAA detect misfolded αSyn via its prion-like seeding activity^23,24^ and have been applied to CSF, brain, and peripheral tissues^25^. These assays offer advantages over immunostaining, including high throughput, reduced interpretive bias, and amplification of trace amounts of aggregation-competent protein^26–28^. While some CSF-based protocols can distinguish PD and MSA, inter-assay reproducibility can be inconsistent, highlighting the need for approaches that combine peripheral accessibility, reliability, and strain-level discrimination.

Cryo-electron microscopy of postmortem brain has shown that αSyn fibril structures differ in MSA and PD^29^. These conformational differences persist in recombinant fibrils templated from patient biospecimens^5^. Furthermore, MSA-derived fibrils exhibit greater seeding potency and neurotoxicity than PD-derived fibrils in cellular systems and mouse models^23,30^. These findings support leveraging structural and biochemical αSyn differences for diagnostic applications using αSyn-SAA^5,6^.

In this cross-sectional, blinded, two-laboratory study, we present a skin-based αSyn-SAA that detects misfolded αSyn in PD and MSA and differentiates them based on aggregation kinetics. The amplified material reveals structural and biochemical properties consistent with disease-specific αSyn strains previously characterized in brain^5^.

## Methods

### Participant recruitment

This study was approved by the ethics committees of the University of Würzburg (#5/19) and Brigham and Women’s Hospital (MGB IRB #2009P000775). Written informed consent was obtained from all participants. Subjects were recruited from the MSA Center of Excellence and MyTrial program, a clinical-trial-ready cohort for neurodegenerative disorders at Brigham and Women’s Hospital (BWH)^31^, and from the Department of Neurology of the University Hospital Würzburg (UKW). Participants included in this study represent a new cohort with respect to skin αSyn-SAA and the data presented here have not been previously published.

We included subjects with a clinically probable or established diagnosis of MSA or PD, and possible or probable Progressive Supranuclear Palsy (PSP) diagnosis, according to the latest MDS criteria for each condition^2,3,32^. Diagnoses were determined by movement disorders specialists. Participants were assessed using the non-motor symptom questionnaire (NMSQ)^33^ and MDS-Unified PD Rating Scale^2^. Healthy controls were age- and sex-matched individuals without evidence of a neurodegenerative disease. Cognitive status was assessed using the Montreal Cognitive Assessment (MoCA). All participants were screened for probable RBD using a single-question screen^34^.

### Skin biopsy procedure and lysate preparation

Skin punch biopsies (3 mm) were obtained from the posterior cervical region (C7), lower back (Th10), lateral thigh, and distal leg (ankle) under local anesthesia. Samples were coded, flash-frozen in liquid nitrogen, and stored at −80°C. Processing and analysis were performed at each site under blinded conditions. Skin lysates (5% w/v) were prepared as previously described^10^ and randomly coded to ensure investigator blinding to diagnosis, clinical data and biopsy site.

### Brain lysates

Post-mortem brainstem tissue (MSA n=3, PD n=3, controls n=3) were obtained from the Brain Bank Center Würzburg, a member of the BrainNet Europe Brain Bank Consortium Network (https://www.brainneteurope.org). Samples were cryopreserved at −80°C immediately after dissection. Lysates (5% w/v in 1% Triton/DPBS) were prepared using a TissueLyser LT (Qiagen), centrifuged at 2,000 × g, and supernatants aliquoted and stored at −80 °C. Brain lysates were diluted 1:100 before αSyn-SAA.

### Skin αSyn-SAA

Two microliters of 10-fold diluted skin homogenate were added to 98 µL of reaction buffer containing 1 mg/mL C-terminus 6xHis-tagged recombinant wild-type αSyn, 500 µM NaCl, 100 mM PIPES, 10 µM thioflavin T (ThT), and 6 ± 2 800 µm silica glass beads (OPS Diagnostics). Substrate was produced at the recombinant protein expression facility of the University of Würzburg^10,11^. Reactions were incubated in clear bottom black 96-well plates (Nunc) at 37°C under cycles of 1 min double orbital shaking at 400 rpm and 5 min rest. The assay ran for 36 hours, and fluorescence was recorded every 45 min at 448/482 nm using a FLUOstar Omega microplate reader (BMG Labtech, Germany). Assay conditions and reader settings were standardized across laboratories, and reproducibility was verified by cross-testing a subset of samples. All available biopsies from each subject were analyzed.

### Kinetic analysis of αSyn-SAA

As illustrated in Fig. 1a, αSyn-SAA positivity was defined using a preestablished fluorescence threshold of 3% of the maximal signal in ≥ 75% of quadruplicate technical replicates. For positive samples, kinetic parameters analyzed included lag time (time to reach fluorescence threshold), Fmax (maximum fluorescence intensity), T50 (time to reach 50% of Fmax), and the area under the curve (AUC). Kinetic analysis was performed on the raw fluorescence data and further refined by fitting each replicate to a sigmoidal function using the Boltzmann method, as described in CSF-based αSyn-SAA studies^35^. Subjects were considered αSyn-SAA positive if at least one biopsy tested positive. To differentiate disease-specific kinetics, a predefined mean Fmax cutoff was applied to distinguish PD-like (Fmax > 50%) from MSA-like (Fmax < 50%) amplification, determined using αSyn-SAA of neuropathologically confirmed PD and MSA cases.

**Figure 1.**
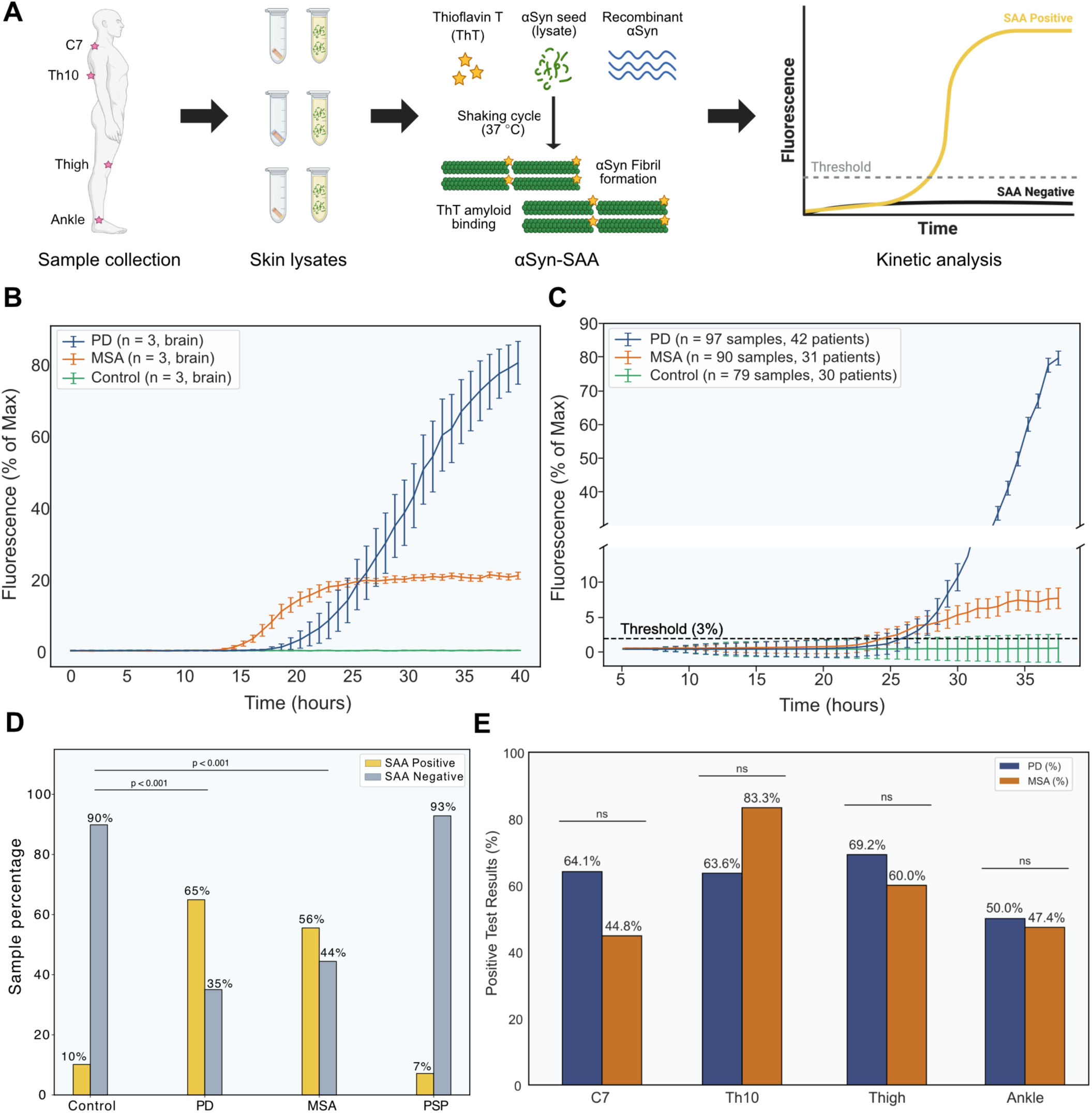
Detection of αSyn seeding activity in skin biopsies. **a.** Overview of the skin αSyn-SAA workflow and anatomical biopsy sites. **b–c.** Median fluorescence curves (±SEM) for αSyn-SAA performed on brain (b) and skin (c) lysates across diagnostic groups; dotted line indicates the fluorescence threshold. **d.** Proportion of positive and negative biopsies per diagnosis (biopsy considered positive when ≥75% of replicates exceeded threshold). **e.** Distribution of positive biopsies across anatomical sites for each diagnostic group.

### Proteinase K treatment and Western blotting

After incubation for 48 hours, αSyn-SAA reaction buffers were collected, pooled, and concentrated using 100 kDa cut-off filters (Amicon). Ten micrograms of aggregated αSyn (quantified by BCA) were digested with 0.12 units of thermolabile Proteinase K (NEB) in Tris buffer (pH 8.0) at 37°C for 1 hour. Reactions were stopped by adding tricine sample buffer and TCEP (Bio-Rad) and heated at 95°C for 10 min. Digested fibrils were resolved on 16% Tris-Tricine gels (Bio-Rad) and transferred to nitrocellulose membranes using an iBlot 2 system (Thermo Fisher). Total αSyn was detected with mouse anti-αSyn (1:2000; BD Biosciences, Cat. # 610786; RRID: AB_398107) and IRDye 800CW secondary antibody (1:10,000; LI-COR Biosciences, 926-32210). The blot was scanned using the Odyssey CLx Infrared Imager.

### Transmission electron microscopy

Fibrils concentrated after αSyn-SAA were examined using transmission electron microscopy (TEM). An aliquot of concentrated fibrillar protein (5 μL of 1 mg/mL) was applied to carbon-coated grids (CF400-CU; Electron Microscopy Sciences) for 10 minutes after rendering the surface hydrophilic through a 20-second glow discharge at 25 mA. Excess liquid was removed using Whatman #1 filter paper, and the grids were briefly floated on a drop of water, blotted, and stained with 1% uranyl acetate (Electron Microscopy Sciences, 22400) for 20–30 seconds. Excess stain was removed with filter paper, and the grids were analyzed using a JEOL 1200EX TEM. Images were captured with an AMT 2k CCD camera, and periodic twist length of fibrils was quantified manually in Fiji.

### Statistical Analysis

Statistical analyses were conducted at both the biopsy and subject levels. Positivity rates across diagnostic groups were compared using Z-tests, and differences across biopsy sites within each group were assessed by Chi-square tests. For positive biopsies, kinetic parameters (lag time, Fmax, T50, AUC) were summarized as mean, median, SD, and SEM, and compared between PD and MSA using Mann–Whitney U-tests due to non-normal distributions. Receiver operating characteristic (ROC) analysis was used to evaluate diagnostic accuracy and to define optimal cutoffs via Youden’s index.

Separate univariate logistic regression models were fitted for PD vs. control and MSA vs. control groups, with αSyn-SAA positivity as the outcome and non-motor symptoms (e.g., RBD, hyposmia, urinary dysfunction) as binary predictors to calculate odds ratios. Mann-Whitney U-tests were used to compare the proportion of positive biopsies between subjects with and without these features. Age and sex differences were tested with one-way ANOVA and Chi-square tests, respectively.

Sensitivity and specificity for αSyn-SAA positivity were calculated at both the biopsy and subject levels by comparing results for each diagnostic group against controls. We evaluated whether aggregation kinetics could discriminate between MSA and PD by assessing concordance between kinetic classification and clinical diagnosis. Kinetic parameters were compared using Mann–Whitney U-tests, and mean fibril twist lengths were analyzed by one-way ANOVA followed by Tukey’s post hoc tests. All tests were two-sided (p < 0.05), with Benjamini–Hochberg correction applied where appropriate. Analyses were performed in Python (v3.12.1) and GraphPad Prism, with figures generated using Matplotlib and finalized in Affinity Designer (Serif).

## Results

### Participants and baseline demographics

This study included 308 skin biopsies from 117 participants: PD (n = 42), MSA (n = 31; 20 parkinsonian [MSA-P], 11 cerebellar [MSA-C]), PSP (n = 14), and 30 cognitively unimpaired controls without neurodegenerative disease. Age and sex did not differ significantly across diagnostic groups (both p > 0.05). Baseline demographics are summarized in Supplementary Table 1. Notably, participants were separate from any prior study.

### Skin αSyn-SAA identifies synucleinopathy with high diagnostic accuracy

Figure 1a illustrates the standardized skin αSyn-SAA workflow implemented across both laboratories. Up to four site-specific biopsies per participant were analyzed under harmonized conditions. Assay thresholds were established using brain lysates from neuropathologically confirmed PD, MSA, and control cases (Fig. 1b), defining both the fluorescence threshold for αSyn-SAA positivity and the maximal fluorescence (Fmax) cutoff that distinguished PD- from MSA-like kinetics. Aggregation profiles obtained across skin biopsies (n = 308; Fig. 1c) closely mirrored the corresponding brain-derived reference curves (Fig. 1b). At the subject level, αSyn-SAA positivity was defined as amplification exceeding the fluorescence threshold in ≥ 75% of replicates from at least one biopsy.

Using these criteria, skin αSyn-SAA detected synucleinopathy with 82.2% sensitivity and 90% specificity versus controls (85.7% vs PSP). Positivity rates were 86% (36/42) in PD and 77% (24/31) in MSA, yielding diagnostic accuracies of 87.5% and 82%, respectively (Table 1). At the biopsy level, 65% (63/97) of PD and 56% (50/90) of MSA samples were positive, both significantly higher than controls (p < 0.0001), whereas PSP did not differ from controls (Fig. 1d). The distribution of positive biopsies did not vary significantly across sites, though thigh samples in PD and lower-back samples in MSA showed the highest positivity (Fig. 1e). Subject- and biopsy-level data are summarized in Table 1.

**Table 1.**
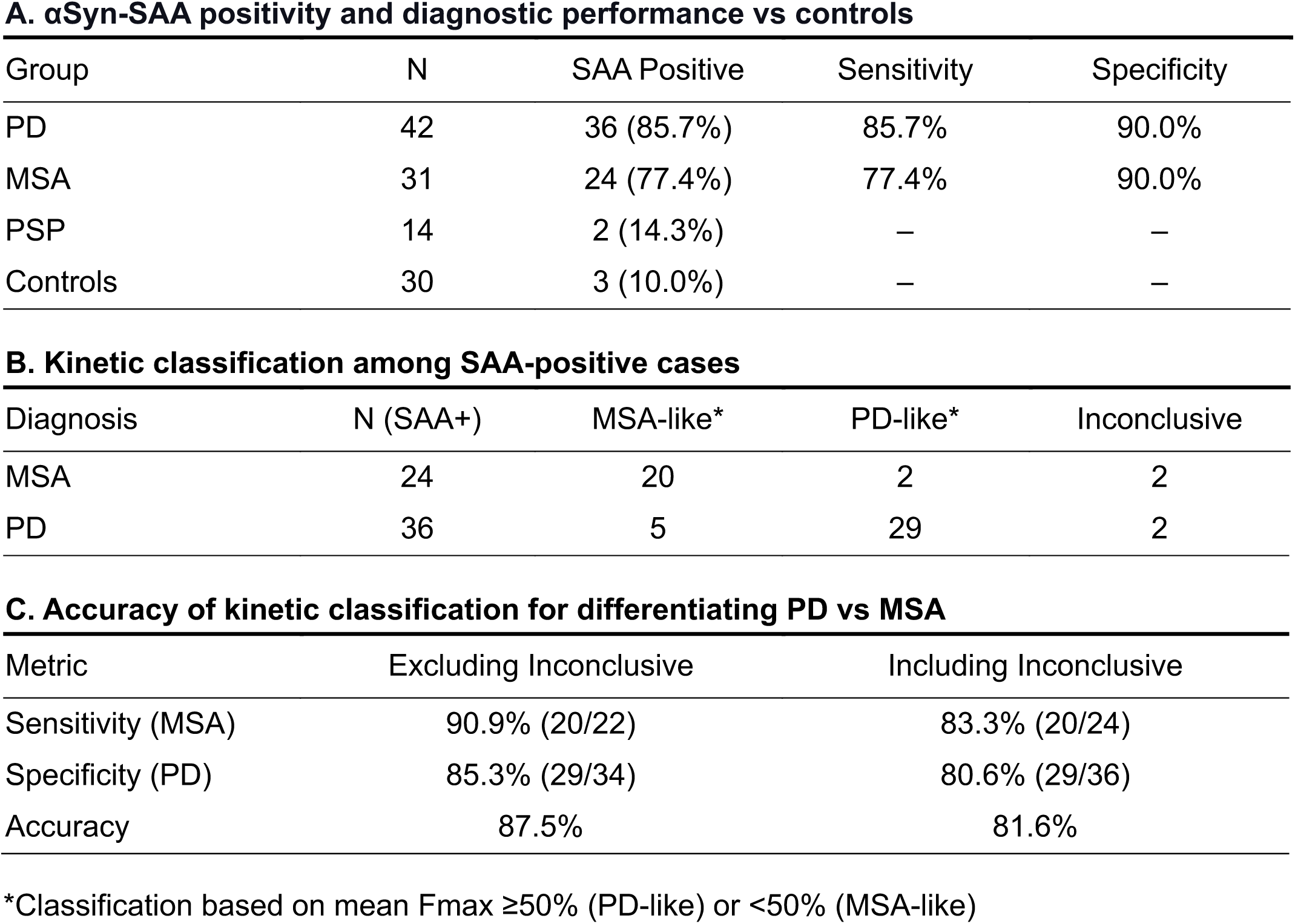
Diagnostic performance of skin αSyn-SAA, including assay positivity, kinetic classification, and accuracy for distinguishing PD from MSA.

**A. αSyn-SAA positivity and diagnostic performance vs controls**
| Group | N | SAA Positive | Sensitivity | Specificity |
| --- | --- | --- | --- | --- |
| PD | 42 | 36 (85.7%) | 85.7% | 90.0% |
| MSA | 31 | 24 (77.4%) | 77.4% | 90.0% |
| PSP | 14 | 2 (14.3%) | — | — |
| Controls | 30 | 3 (10.0%) | — | — |

**B. Kinetic classification among SAA-positive cases**
| Diagnosis | N (SAA+) | MSA-like* | PD-like* | Inconclusive |
| --- | --- | --- | --- | --- |
| MSA | 24 | 20 | 2 | 2 |
| PD | 36 | 5 | 29 | 2 |

**C. Accuracy of kinetic classification for differentiating PD vs MSA**
| Metric | Excluding Inconclusive | Including Inconclusive |
| --- | --- | --- |
| Sensitivity (MSA) | 90.9% (20/22) | 83.3% (20/24) |
| Specificity (PD) | 85.3% (29/34) | 80.6% (29/36) |
| Accuracy | 87.5% | 81.6% |

\*Classification based on mean Fmax ≥ 50% (PD-like) or <50% (MSA-like)

Two PSP patients showed αSyn-SAA positivity despite clinical diagnoses of probable PSP-Richardson’s syndrome and possible PSP with corticobasal features, both exhibiting akinetic-rigid, levodopa-resistant parkinsonism without non-motor symptoms suggestive of synucleinopathy^32^. Three healthy controls were also αSyn-SAA positive, each with at least two positive biopsies. One reported urinary urgency and constipation, while the others had no features of prodromal synucleinopathy^36^.

### Kinetic analysis of skin αSyn-SAA differentiates MSA from PD

Based on αSyn-SAA aggregation kinetics in brain, a mean Fmax threshold of 50% was used to classify skin results as either *PD-like* (FMax > 50%) or *MSA-like* (FMax < 50%) (Fig. 1b). Kinetic profiles were first assigned at the biopsy level and then aggregated per subject based on the predominant pattern. Subjects with equal numbers of *PD-* and *MSA-like* biopsies were considered inconclusive. Among 60 subjects with αSyn-SAA-positive synucleinopathy (PD = 36, MSA = 24), 29 PD cases exhibited *PD-like* profiles, while 20 MSA cases showed *MSA-like* kinetics (Fig. 2a). Seven subjects (11.7%) demonstrated discordance with the clinical diagnosis and four (6.7%) were inconclusive. Overall, concordance with a clinical diagnosis among αSyn-SAA-positive subjects was 81.6% (Table 1).

**Figure 2.**
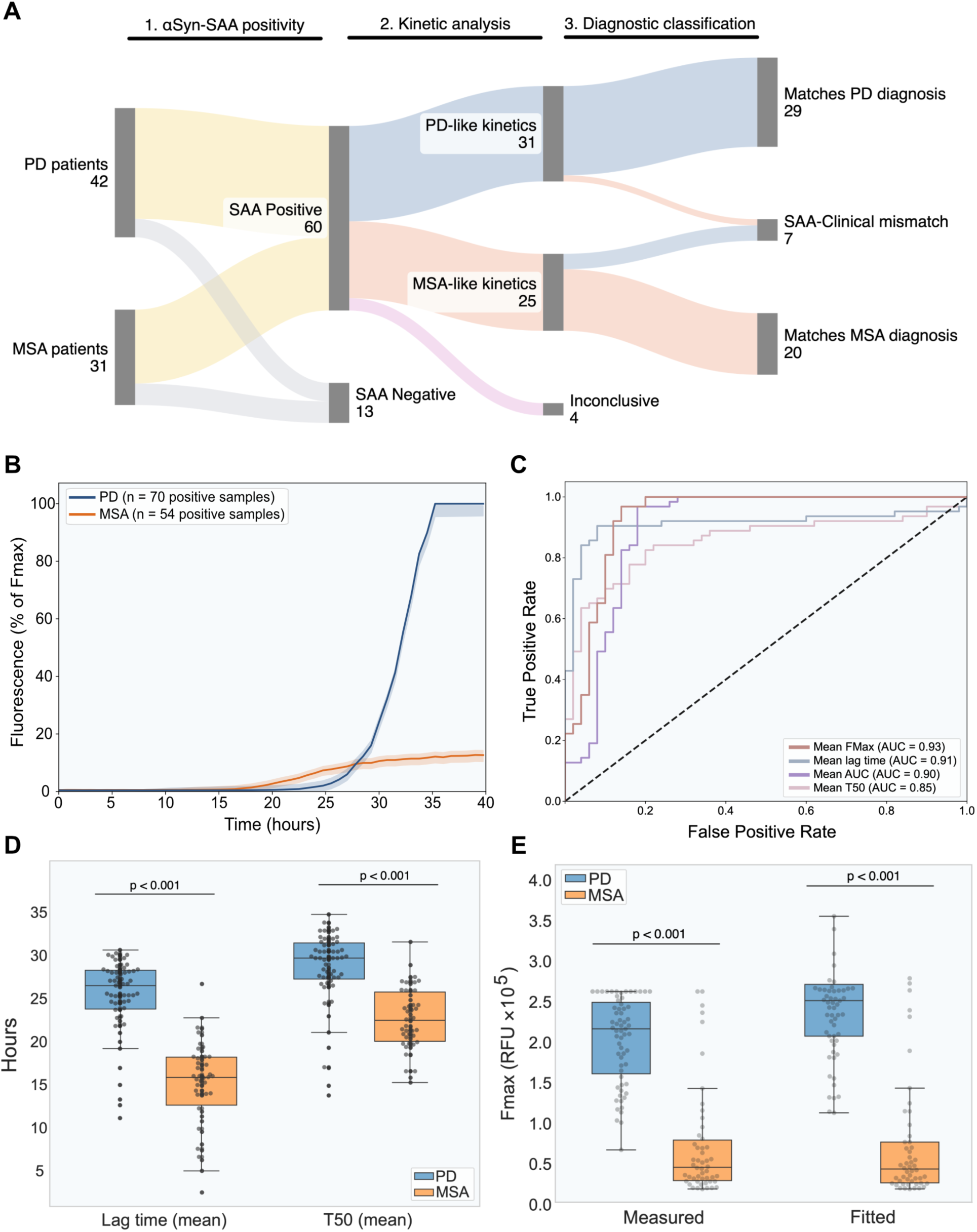
Kinetic analysis of skin αSyn-SAA discriminates between PD and MSA. **a.** Classification of subjects based on αSyn-SAA positivity, kinetic profile (PD-like vs MSA-like), and concordance with clinical diagnosis. **b.** Median fluorescence curves (±SEM) for positive αSyn-SAA samples in PD and MSA. **c.** ROC curves for kinetic parameters (Fmax, T50, lag time, AUC) differentiating PD and MSA. **d.** Distribution of the mean lag time and T50 across positive biopsies, with each dot representing an individual sample. **e.** Maximal fluorescence (Fmax) across diagnoses, shown for measured and sigmoidal-fitted values.

At the biopsy level, kinetic profiling revealed distinct amplification signatures differentiating PD and MSA (Fig. 2b). ROC analysis demonstrated excellent discrimination among αSyn-SAA positive samples (AUC = 0.93 for mean Fmax; 0.91 for mean lag time; Fig. 2c). PD samples exhibited significantly higher Fmax (77.06 ± 20.4%), than MSA (27 ± 26%; p < 0.0001), whereas MSA aggregated faster, with shorter lag times (14.8 ± 4.9 h vs 25.1 ± 4.4 h, p < 0.0001) and T50 values (Fig. 2d; Supplementary table 2). These kinetic distinctions were consistent across all biopsy sites. Sigmoidal curve fitting, consistent with established CSF αSyn-SAA methods^35^, reproduced the differences across predicted Fmax values (Fig. 2e). The optimal Fmax cutoff from ROC analysis (Fmax = 49.4%) closely matched the predefined 50% threshold derived from brain assays.

### αSyn seeding activity correlates with key non-motor features in MSA and PD

Neuropathological confirmation remains the gold standard for diagnosing synucleinopathies but is rarely available in clinical research. In its absence, clinical features that reflect underlying αSyn pathology offer a means to validate peripheral biomarker findings. We therefore evaluated whether skin αSyn-SAA positivity correlates with core non-motor features of PD and MSA. Some of these features, such as RBD, are shared across synucleinopathies, whereas others, including hyposmia, are disease-specific and incorporated into current diagnostic criteria^3,32^. Logistic regression analyses were performed using αSyn-SAA positivity as the dependent variable and non-motor symptoms as predictors, modeled separately for PD and MSA versus controls.

Among MSA cases, urinary urgency or urge incontinence, core autonomic features of the 2022 MSA diagnostic criteria^3^, were strongly associated with αSyn-SAA positivity (OR: 19.59, 95% CI: 3.74–102.5), whereas no such association was observed in PD. In contrast, hyposmia, one of the earliest and most characteristic non-motor features of PD^2,36^, demonstrated a positive but non-significant trend (OR 2.6, 95% CI 0.96–6.9). RBD, a well-established prodromal feature of synucleinopathy^2,3,36^, was significantly associated with αSyn-SAA positivity in both MSA (OR 4.3, 95% CI 1.35–13.6) and PD (OR 5.6, 95% CI 1.9–16.2; Fig. 3a).

**Figure 3.**
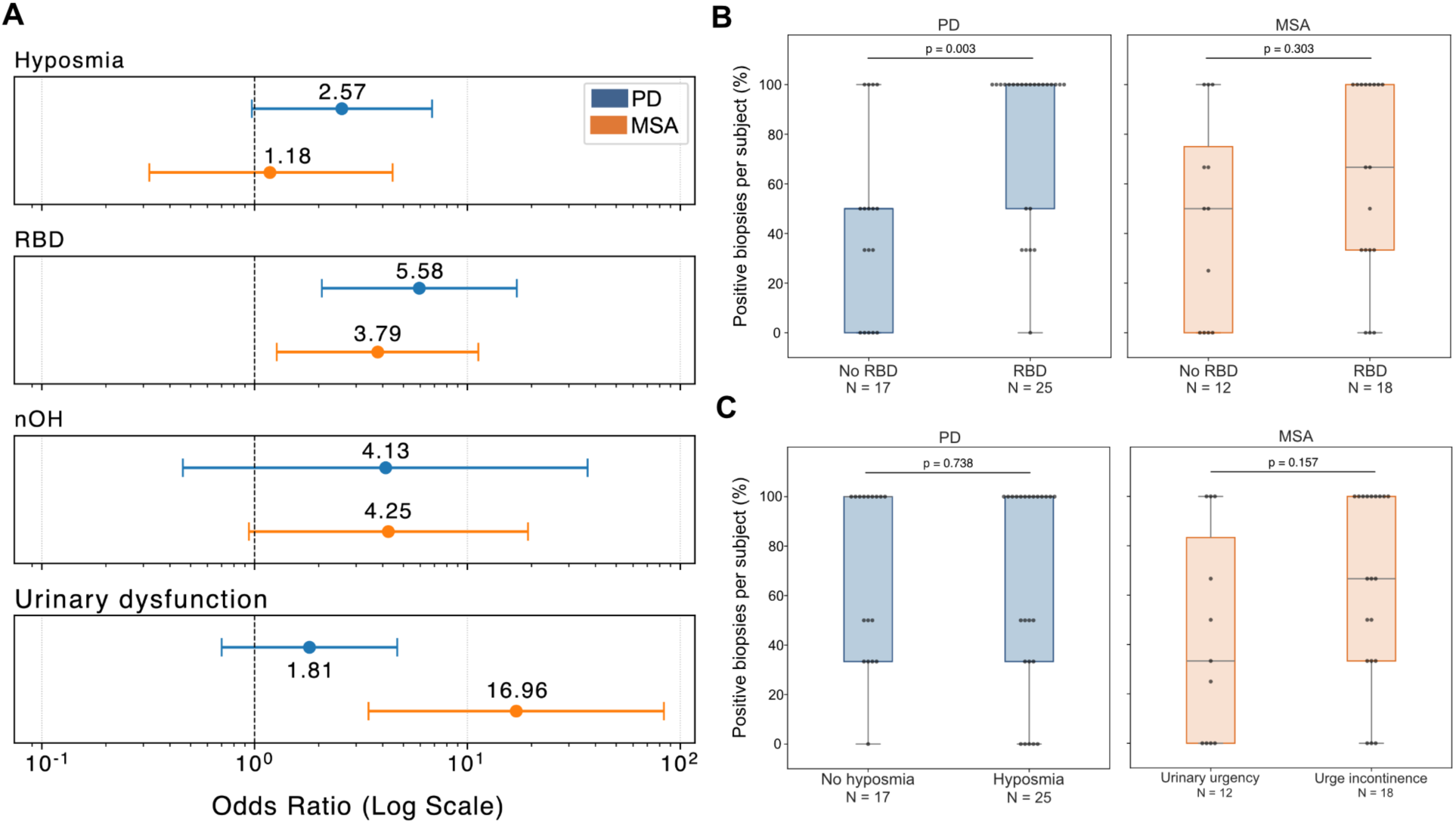
Associations between non-motor features and αSyn-SAA positivity. **a.** Odds ratios (95% CI) from logistic regression models linking non-motor symptoms with αSyn-SAA positivity in PD and MSA. **b.** Proportion of positive biopsies per subject stratified by RBD status in PD (left) and MSA (right). **c.** Proportion of positive biopsies stratified by hyposmia in PD (left) and urge incontinence in MSA (right). RBD: REM sleep behavior disorder. nOH: neurogenic orthostatic hypotension.

### Subjects with RBD are more likely to exhibit αSyn-SAA positivity in multiple biopsies

We next assessed whether the pathological burden, estimated by the proportion of αSyn-SAA-positive biopsies per subject, correlated with clinical phenotypes. In PD, RBD was significantly associated with a higher burden of peripheral αSyn pathology (p = 0.003, Fig. 3b, left). 18 out of 25 PD patients with RBD showed αSyn-SAA positivity across all biopsy sites. A similar, though non-significant trend was observed among MSA cases (Fig. 3b, right), whereas no such pattern was seen with hyposmia (Fig. 3c, left). Among MSA subjects, those with urge incontinence or neurogenic orthostatic hypotension also exhibited higher proportions of positive biopsies, though these associations did not reach significance, likely due to limited sample size (Fig. 3c, right). No correlations were observed between αSyn-SAA positivity and disease duration, diagnostic certainty, or MSA subtype.

### Alpha-synuclein strain differences between PD and MSA are reproduced in skin biopsies

As noted above, aggregation kinetics from brain samples (n = 3 per group) closely matched those observed in skin (n = 308) under standardized αSyn-SAA conditions (Fig. 1). The difference in endpoint fluorescence between MSA- and PD-seeded reactions was sufficiently large to produce distinct color changes visible to the naked eye (Fig. 4a). The yield of fibrillar αSyn did not significantly differ by diagnosis or tissue type: mean ± SD values were 59.6 ± 18.2 µg for PD skin (n = 5), 60.1 ± 16.7 µg for MSA skin (n = 5), 60.0 ± 17.3 µg for PD brain (n = 3), and 64.1 ± 15.8 µg for MSA brain (n = 3), based on quadruplicate reactions. Two-way ANOVA revealed no significant main effects of diagnosis or tissue.

**Figure 4.**
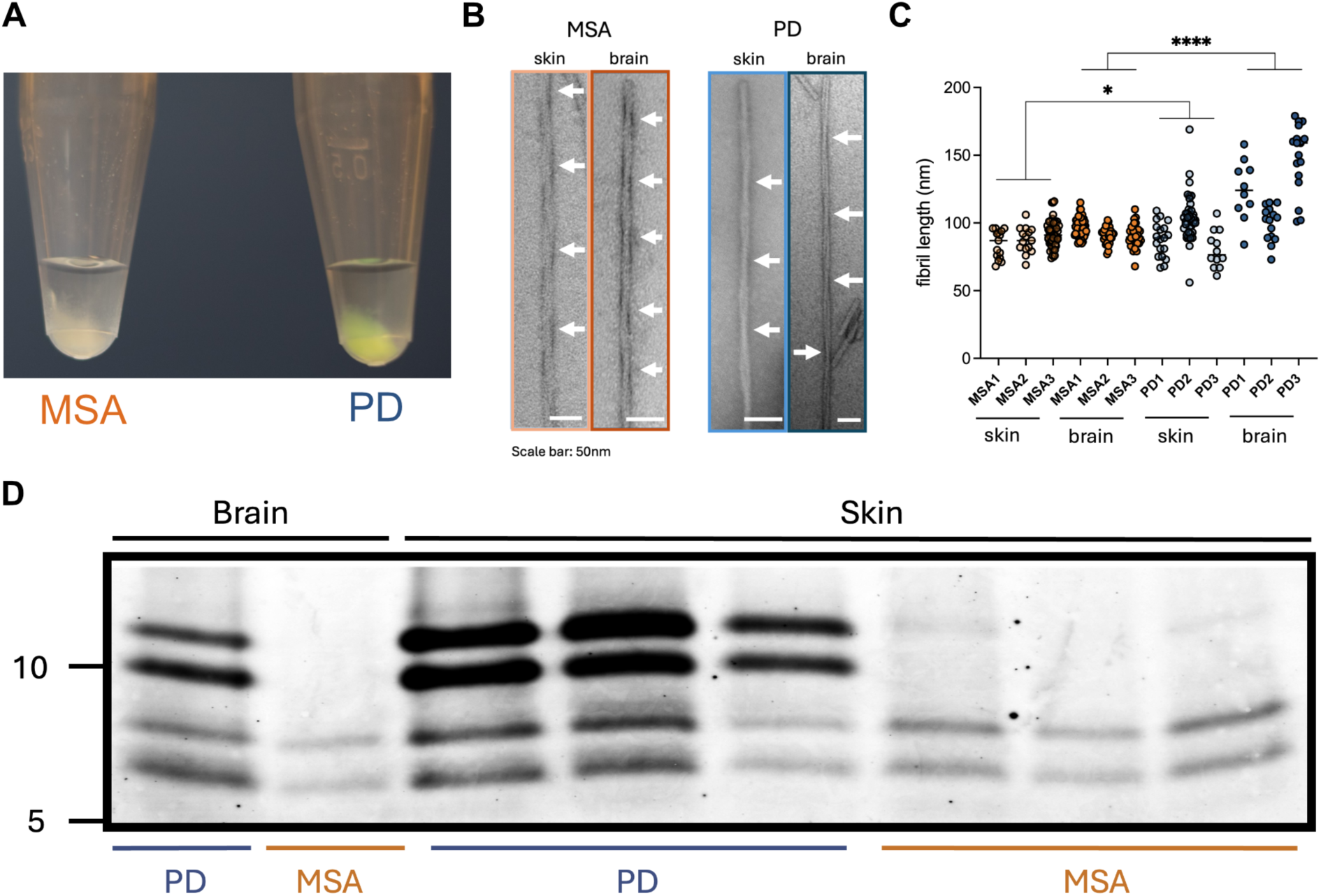
Disease-specific fibril properties in αSyn-SAA–amplified products from skin and brain. **a.** Representative photograph of the fibril-enriched buffer collected after αSyn-SAA seeded with MSA (appearing white) or PD (yellow) samples. **b.** TEM images showing morphological differences in PD- and MSA-derived fibrils (scale bar: 50 nm). **c.** Quantification of the periodic spacing in fibrils derived from skin biopsies and postmortem tissue of patients with PD and MSA (n = 3 per group, **** = p < 0.001). **d.** Proteinase K digestion profiles of αSyn-SAA–amplified brain and skin lysates (n = 3 per disease group).

Negative-stain TEM revealed consistent ultrastructural differences between strains: PD-amplified fibrils exhibited longer inter-helical crossover distances than MSA fibrils, both in brain-and skin-derived material (p < 0.0001 and p = 0.026, respectively; Fig. 4b,c). Proteinase K fingerprinting demonstrated distinct digestion patterns, showing a characteristic four-band pattern in PD (∼5–10 kDa) and only the two lower molecular weight bands in MSA (Fig. 4d). These proteolytic signatures were conserved across brain- and skin-derived fibrils within each diagnosis, consistent with prior CSF findings^5^.

In a parallel cohort of early clinically diagnosed MSA cases, we integrated skin αSyn-SAA with [¹⁸F]ACI-12589 PET to strengthen the biological diagnosis of early MSA^37^. In a neuropathologically confirmed case from this cohort, αSyn-SAA was applied across skin, brain, and CSF from the same subject, together with postmortem autoradiography and pS129-αSyn immunostaining. αSyn-SAA revealed highly concordant aggregation kinetics across tissues, suggesting that MSA-specific αSyn strain properties are preserved across tissues under uniform assay conditions^37^.

## Discussion

The development of αSyn-SAA has transformed the detection of misfolded αSyn across biospecimens, an important step in the biological characterization of synucleinopathies^5–8,10,11,15^. Aggregates amplified from brain and CSF in PD and MSA exhibit distinct kinetic, biochemical, and ultrastructural signatures—hallmarks of disease-specific αSyn strains^5^. While CSF-based assays are increasingly validated^25^, there is growing interest for less invasive approaches, including skin biopsies. A key question is whether αSyn aggregates detected peripherally retain the structural fidelity of brain-derived species and whether such assays can reliably distinguish among synucleinopathies in clinical cohorts.

In this two-laboratory study, skin αSyn-SAA revealed distinct aggregation kinetics that differentiated MSA from PD across more than 300 biopsies, reproducing the kinetic and structural hallmarks of disease-specific αSyn strains previously identified in brain and CSF^5^. In this same assay, brain homogenates from pathology-confirmed cases showed analogous kinetic profiles: slower aggregation with higher fluorescence in PD and faster, lower-intensity aggregation in MSA. Proteinase K digestion and TEM of amplified material confirmed that skin-derived aggregates preserve strain-specific fibril morphologies^5^. In related work, application of this αSyn-SAA to matched skin, CSF, and brain samples from a neuropathologically confirmed MSA case revealed consistent kinetic signatures across all tissues^37^. Collectively, these findings support the ability of skin αSyn-SAA to capture disease-relevant strains reflective of central nervous system pathology.

Skin αSyn-SAA identified synucleinopathy in 86% of PD and 77% of MSA cases, while remaining negative in 90% of healthy controls and 85.7% of PSP cases. These findings align with our prior smaller studies and with independent reports demonstrating detectable αSyn seeding activity in skin (sensitivity, 77–93%; specificity, 87–100%)^10,11,15^ and approach the pooled performance of CSF αSyn-SAA (88% sensitivity, 95% specificity)^25^. The present work substantially extends these observations by including a larger blinded cohort and enabling direct PD–MSA comparisons within a unified assay framework.

Among αSyn-SAA-positive biopsies, ROC analysis of kinetic parameters showed excellent discrimination between MSA and PD strains. To operationalize this distinction for differential diagnosis, αSyn-SAA–positive samples were classified as *MSA-like* or *PD-like* based on an Fmax threshold derived from pathology-confirmed brain. Using this criterion, 83% of clinically diagnosed MSA cases were classified as MSA-like and 81% of PD cases as PD-like, yielding overall concordance with clinical diagnosis in most αSyn-SAA–positive subjects. Although our PSP sample was small, two PSP cases showed αSyn-SAA positivity, consistent with emerging evidence of mixed αSyn co-pathology in PSP^38^.

A small subset showed discordance between kinetic classification and clinical diagnosis (2 PD and 5 MSA), while four additional cases (2 PD, 2 MSA) were inconclusive owing to divergent results across biopsies. As illustrated in parallel work, such discordances may reflect misdiagnoses or early phenotypes with overlapping clinical features^37^. For instance, PD with prominent neurogenic orthostatic hypotension that can initially meet criteria for probable MSA. Longitudinal follow-up will be essential to determine whether early αSyn-SAA signatures anticipate eventual clinical reclassification. In the context of emerging disease-modifying trials, biomarker results that diverge from the clinical impression should prompt careful reevaluation to improve diagnostic precision and participant stratification in research settings.

A key limitation of this study is the absence of neuropathological confirmation, which precludes definitive assessment of diagnostic accuracy. As a surrogate, we examined whether characteristic non-motor features of each synucleinopathy were associated with αSyn-SAA positivity. In both PD and MSA, a history of RBD was associated with a two- to threefold higher likelihood of a positive result. Within MSA, neurogenic orthostatic hypotension and urinary dysfunction were each independently associated with αSyn-SAA positivity, whereas in PD, self-reported hyposmia showed the strongest but non-significant trend. No associations were found with disease duration, echoing prior CSF studies and underscoring that conventional αSyn-SAA remains a qualitative assay better suited for diagnosis than for disease staging^5,10,11^. The ongoing development of quantitative or digital αSyn-SAA may help address this limitation^26^.

Unlike studies employing αSyn immunofluorescence (αSyn-IF)^19–22,39–41^, we observed no site-specific differences in αSyn-SAA positivity, consistent with our earlier observations using the same platform^10,11^. This may reflect the high (femtogram-level) sensitivity of SAA to αSyn fibrils^26^. PD subjects with RBD were more likely to exhibit αSyn-SAA positivity across multiple biopsy sites compared with those without RBD, whereas hyposmia was not associated with a more diffuse pattern of positivity. This discrepancy may suggest that skin αSyn-SAA preferentially detects cases with more widespread or peripherally distributed αSyn pathology, contrasting with CSF-based assays, where hyposmia has been strongly linked to αSyn-SAA positivity^4^. It is therefore possible that αSyn distribution varies by phenotype, such that skin and CSF αSyn-SAA capture distinct aspects of disease biology; a hypothesis that warrants evaluation. Olfactory function in this cohort was self-reported, a method known to underestimate the true prevalence of olfactory impairment. We are now incorporating quantitative smell testing alongside skin αSyn-SAA in ongoing work to validate this finding. Among MSA subjects, the presence of RBD, urge incontinence, or neurogenic orthostatic hypotension also trended toward higher αSyn-SAA burden, although these associations did not reach statistical significance.

Direct comparisons between skin αSyn-IF and αSyn-SAA are challenging due to key methodological differences. Although prior work suggests substantial overlap between the two techniques, each may detect cases that the other misses^27^. Phosphorylated αSyn detected by αSyn-IF is a robust marker of Lewy pathology but it may encompass a broad range of conformers, including species that may not seed aggregation. Conversely, αSyn-SAA selectively amplifies aggregation-competent species and may detect pathology in sparse or patchy inclusions that fall below the αSyn-IF threshold, albeit without providing information on αSyn distribution [refs]. The biological distinction between seed-competent and seed-incompetent aggregates has been recapitulated in our induced pluripotent stem cell (iPSC)-derived “inclusionopathy” models and supported by recent observations in LRRK2-associated PD^4,42^. Consistent with this, we have identified cases with αSyn-SAA positivity despite negative αSyn-IF staining (data not shown), underscoring the complementary nature of these assays.

Although skin sampling is less invasive than lumbar puncture, our protocol required multiple biopsies per participant, consistent with αSyn-IF paradigms increasingly used in clinical practice. The diagnostic performance of a single biopsy, and the optimal site, remain under investigation. Direct comparison of skin and CSF αSyn-SAA remains an outstanding question; preliminary experience suggests that each may capture distinct aspects of αSyn biology. Ongoing studies in our group are acquiring paired skin–CSF samples to address this gap.

The skin biopsy platform also enables patient-specific translational applications. Parallel assessment of αSyn pathology by αSyn-IF and SAA is ongoing at our centers, and fibroblasts derived from biopsies collected at the same visit can be reprogrammed into iPSCs^31^. These patient-derived neuronal and glial models can then be seeded with patient-amplified αSyn to recapitulate disease-relevant pathology^52^, providing a versatile system for mechanistic studies and screening αSyn-targeting diagnostics and therapies, including PET radiotracers and candidate disease-modifying compounds.

In sum, with its high-throughput format, scalability, and semi-automated readout, skin αSyn-SAA expands the diagnostic and research potential of the skin biopsy platform. It complements αSyn-IF-based methods while overcoming limitations related to sample throughput and analytic subjectivity. The assay’s exceptional sensitivity enables multiple experimental runs from a single specimen and supports longitudinal sampling, multicenter implementation, and assay standardization. Skin αSyn-SAA thus provides a less invasive and scalable biomarker with diagnostic performance comparable to CSF-based assays. While a robust plasma-based αSyn-SAA assay is eagerly awaited, the ability to combine tissue-level localization with conformational strain analysis remains a unique strength of the skin approach. By capturing disease-specific aggregation kinetics, skin αSyn-SAA enables molecular distinction between PD and MSA, offering a practical tool for clinical stratification and for refining the biological definition of synucleinopathies^43,44^.

## Data Availability

All data produced in the present work are contained in the manuscript. Raw fluorescence data from αSyn-SAA are available upon request to the authors

## Acknowledgments

We are deeply grateful to the study participants for contributing samples. We thank Dr. Lars Schönemann from the recombinant protein expression facility of the Rudolf–Virchow Center for the expression and purification of α-synuclein, and Maria Ericsson and the team at the Harvard Medical School Electron Microscopy Facility for technical support.

## Authors’ roles

A.K., D.R., R.B., C.M., C.S., B.C., V.K., and K.D. contributed to the conception and design of the study.

A.K., D.R., C.P., A.N., A.K.-2, S.R., K.S., L. H.-R., O.L., J.M., S.P., K.J., D.C., S.K., performed acquisition and analysis of data.

A.K., D.R., A.N., V.K., and K.D. contributed to drafting the manuscript and/or figures.

---

**Supplementary Table 1.** Participant demographics and clinical features by diagnostic group.

|  | PD<br>(n = 42) | MSA<br>(n = 31) | PSP<br>(n = 14) | Controls<br>(n = 30) |
| --- | --- | --- | --- | --- |
| Age in years, mean (SD) | 67.6 (8.84) | 64.0 (14.2) | 68.7 (5.2) | 62.3 (10.7) |
| Sex |  |  |  |  |
| Female (%) | 13 (31%) | 15 (48%) | 10 (71%) | 12 (40%) |
| Male (%) | 29 (69%) | 16 (52%) | 4 (29%) | 18 (60%) |
| Diagnostic certainty |  |  |  |  |
| Established PD | 31 (74%) | -- | -- | -- |
| Probable PD | 11 (26%) | -- | -- | -- |
| Established MSA | -- | 8 (26%) | -- | -- |
| Probable MSA | -- | 23 (74%) | -- | -- |
| Probable PSP | -- | -- | 8 (57%) | -- |
| Possible/Suggestive PSP | -- | -- | 6 (43%) | -- |
| MSA subtype |  |  |  |  |
| Cerebellar (MSA-C) | -- | 11 (35%) | -- | -- |
| Parkinsonian (MSA-P) | -- | 20 (65%) | -- | -- |
| Stage (Hoehn and Yahr), mean (SD) | 2.6 (0.8) | 2.9 (1.2) | 3.4 (0.7) | -- |
| Time since diagnosis (years), mean (SD) | 8 (6.4) | 2.35 (1.9) | 2.21 (2.6) | -- |
| RBD | 25 (60%) | 18 (60%) | 1 (7%) | 6 (20%) |
| Orthostatic hypotension | 12 (29%) | 21 (68%) | 4 (29%) | 0 (0%) |
| Constipation | 17 (40%) | 27 (93%) | 3 (21%) | 5 (17%) |
| Hyposmia | 25 (60%) | 5 (18%) | 3 (21%) | 6 (20%) |
| Urinary dysfunction |  |  |  |  |
| Urgency or frequency | 17 (40%) | 11 (35%) | 6 (43%) | 6 (20%) |
| Urge incontinence | 9 (21%) | 20 (65%) | 2 (14%) | 2 (7%) |

**Supplementary Table 2.** Kinetic parameters of skin αSyn-SAA. Positivity rates and mean kinetic parameters (Fmax, lag time, T50) for all biopsies per diagnostic group.

| Biopsy-level kinetic analysis |  |  |  |  |
| --- | --- | --- | --- | --- |
|  | PD (n = 97) | MSA (n = 90) | PSP (n = 42) | HC (n = 79) |
| Total |  |  |  |  |
| Positive (% of total) | 63 (64.9%) | 50 (55.6%) | 3 (7.1%) | 8 (10.1%) |
| Mean Fmax, % (SD) | 77.06 (20.4) | 26.90 (26.2) | 77.86 (26.4) | 56.30 (34.2) |
| Mean Lag time, h (SD) | 25.07 (4.4) | 14.81 (4.9) | 22.70 (7.0) | 15.80 (4.9) |
| Mean T50, h (SD) | 28.18 (4.5) | 22.34 (3.7) | 24.63 (6.1) | 20.17 (5.5) |
| C7 |  |  |  |  |
| Positive (% of total) | 25 (64%, 39) | 13 (45%, 29) | 1 (7%, 14) | 3 (11%, 28) |
| Mean Fmax, % (SD) | 78.20 (14.8) | 31.23 (35.5) | 48.60 | 70.17 (35.1) |
| Mean Lag time, h (SD) | 25.69 (3.7) | 16.35 (5.1) | 18.96 | 15.38 (6.1) |
| Mean T50, h (SD) | 28.87 (3.3) | 24.3 (3.9) | 22.50 | 19.38 (6.8) |
| Th10 |  |  |  |  |
| Positive (% of total) | 7 (64%, 11) | 10 (83%, 12) | 0 (0%, 12) | 3 (13%, 24) |
| Mean Fmax, % (SD) | 81.80 (25.1) | 21.87 (9.0) | -- | 42.72 (18.8) |
| Mean Lag time, h (SD) | 26.93 (1.7) | 17.15 (2.8) | -- | 18.47 (4.8) |
| Mean T50, h (SD) | 29.76 (1.6) | 21.58 (4.0) | -- | 23.09 (3.8) |
| Thigh |  |  |  |  |
| Positive (% of total) | 27 (69%, 39) | 18 (60%, 30) | 2 (15%, 13) | 2 (9%, 23) |
| Mean Fmax, % (SD) | 75.16 (24.2) | 23.16 (23.3) | 92.49 (10.6) | 55.88 (62.4) |
| Mean Lag time, h (SD) | 23.93 (5.4) | 13.46 (4.9) | 24.56 (8.8) | 12.47 (1.5) |
| Mean T50, h (SD) | 26.99 (5.9) | 22.09 (2.9) | 25.69 (8.2) | 16.97 (6.2) |
| Ankle |  |  |  |  |
| Positive (% of total) | 4 (50%, 8) | 9 (47%, 19) | 0 (0%, 3) | 0 (0%, 4) |
| Mean Fmax, % (SD) | 74.46 (21.7) | 33.72 (30.16) | -- | -- |
| Mean Lag time, h (SD) | 25.64 (2.6) | 12.72 (5.3) | -- | -- |
| Mean T50, h (SD) | 29.25 (2.2) | 20.88 (3.6) | -- | -- |

